## Supplementary figures and images for "Healthcare utilization and expenditure among people with type 2 diabetes and/or hypertension in Cambodia: results from a cross-sectional survey"

### Annexure 1

**Annexure 1. Locations of the five studied ODs in Cambodia, 2020.**

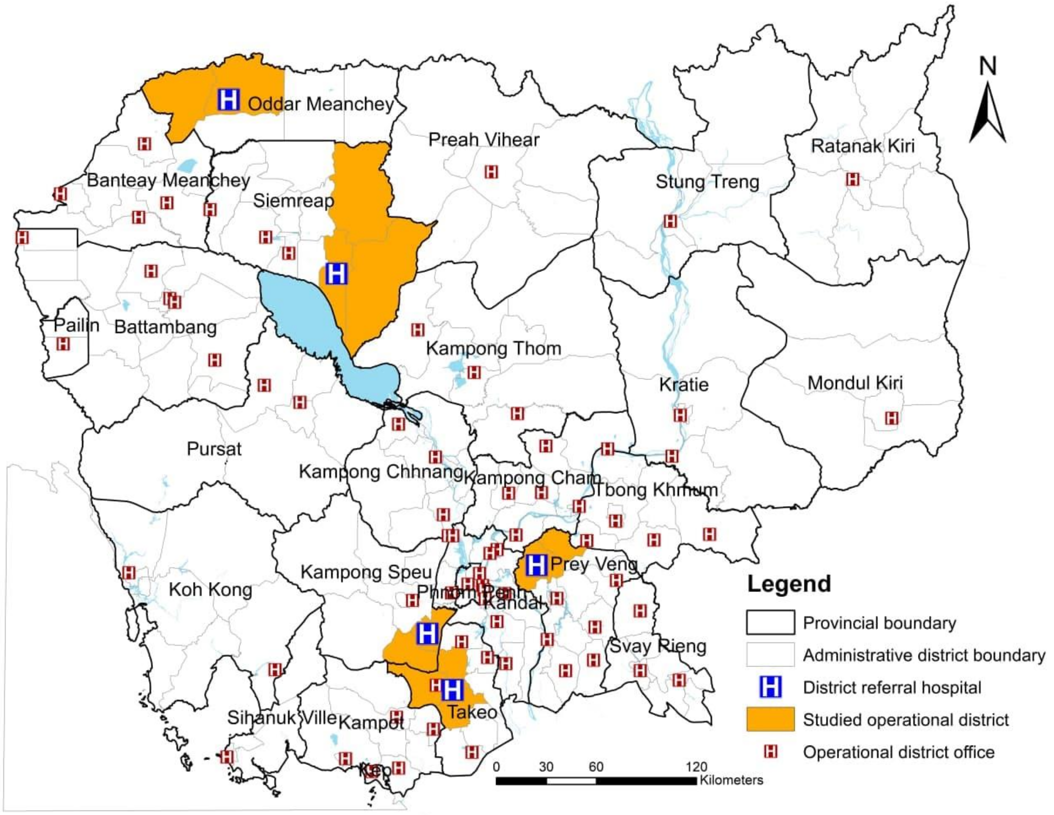
