## Supplementary material for "Healthcare utilization and expenditure among people with type 2 diabetes and/or hypertension in Cambodia: results from a cross-sectional survey": Annexure 2

### Annexure 2. Principal Component Analysis

The wealth quintile was used to classify participants into five socio-economic classes: poorest, poor, medium, rich, and richest. The wealth quintile was constructed using principal component analysis (PCA). The head of the family or family representative was interviewed using a 30-item tool that was taken from the 2014 Cambodia Demographic Health Survey. The tool is designed to classify households' wealth based on their properties. The binary answer questions were coded with 0 for no and 1 for yes. The questions with non-binary-answer were split into binary questions with a code of 0 for no and 1 for yes [21]. The first score component was used to classify the wealth into five quintiles [21]. The lower score represents the poorer status, and the higher score represents the richer status. The list of questions used for PCA is in the table below.

### HOUSEHOLD SOCIO-ECONOMIC STATUS

| ***QH3. Does your household have? [CDHS]*** | | |
|---|---|---|
| 301. electricity | 1=Yes | 0=No |
| 302. a radio? | 1=Yes | 0=No |
| 303. a television? | 1=Yes | 0=No |
| 304. a mobile telephone? | 1=Yes | 0=No |
| 305. If yes, is it a smartphone? | 1=Yes | 0=No |
| 306. a refrigerator? | 1=Yes | 0=No |
| 307. a wardrobe? | 1=Yes | 0=No |
| 308. a sewing machine or loom? | 1=Yes | 0=No |
| 309. CD/DVD player? | 1=Yes | 0=No |
| 310. a generator/battery/solar panel? | 1=Yes | 0=No |

| Does any member of your household own/have ? [CDHS] | | |
|---|---|---|
| 311. a watch? | 1=Yes | 0=No |

| | | |
|---|---|---|
| 312. a bicycle or cyclo? | 1=Yes | 0=No |
| 313. a motorcycle or motor-scooter? | 1=Yes | 0=No |
| 314. a motorcycle-cart | 1=Yes | 0=No |
| 315. a oxcart or horsecart? | 1=Yes | 0=No |
| 316. a car or truck, tractor or van? | 1=Yes | 0=No |
| 317. a boat with a motor? | 1=Yes | 0=No |
| 318. a boat without a motor? | 1=Yes | 0=No |
| 319. any agricultural land? | 1=Yes | 0=No |
| 320. any livestock, herds, other farm animals, or poultry? | 1=Yes | 0=No |
| 321. a bank account? | 1=Yes | 0=No |

| | | |
|---|---|---|
| 322 | What is the main material of the floor?<br><br>*Record observation* | 1. Earth/sand/clay<br>2. Dung<br>3. Wood planks<br>4. Palm/bamboo<br>5. Parquet or polished wood<br>6. Vinyl or Asphalt strips<br>7. Ceramic tiles<br>8. Cement tiles<br>9. Cement<br>10. Floating house<br>11. Other (specify) |
| 323 | What is the main material of the roof?<br><br>*Record observation* | 1. Bamboo/thatch/palm leaf<br>2. Rustic mat<br>3. Wood planks<br>4. Cardboard<br>5. Plastic sheet<br>6. Metal<br>7. Wood<br>8. Calamine/cement fiber<br>9. Ceramic tiles<br>10. Clay tiles<br>11. Cement<br>12. Other (specify) |
| 324 | What is the main material of the exterior walls?<br><br>*Record observation* | 1. Palm/bamboo/thach<br>2. Dirt<br>3. Bamboo with mud<br>4. Straw with mud<br>5. Stone with mud<br>6. Uncovered adobe |

| | | |
|---|---|---|
| | | 7. Plywood<br>8. Cardboard<br>9. Reused wood<br>10. Metal<br>11. Cement<br>12. Stone with lime/cement<br>13. Bricks<br>14. Cement blocks<br>15. Covered adobe<br>16. Wood planks/shingles<br>17. Other (specify) |
| 325 | How many rooms in this household are used for sleeping? | Rooms:………………… |
| 326 | What is the main source of drinking water during the wet season for members of your household? | 1. Piped into dwelling<br>2. Piped to yard/plot<br>3. Public tap/standpipe<br>4. Tube well or borehole<br>5. Protected well<br>6. Unprotected well<br>7. Protected spring<br>8. Unprotected spring<br>9. Rainwater<br>10. Tanker truck<br>11. Cart with small tank<br>12. Surface water (river/dam/lake/pond/stream/canal/irrigation channel<br>13. Bottled water<br>14. Other (specify) |
| 327 | What is the main source of drinking water during the dry season for members of your household? | 1. Piped into dwelling<br>2. Piped to yard/plot<br>3. Public tap/standpipe<br>4. Tube well or borehole<br>5. Protected well<br>6. Unprotected well<br>7. Protected spring<br>8. Unprotected spring<br>9. Rainwater<br>10. Tanker truck<br>11. Cart with small tank<br>12. Surface water (river/dam/lake/pond/stream/canal/irrigation channel<br>13. Bottled water<br>14. Other (specify) |

| | | |
|---|---|---|
| 328 | Do you do anything to the water to make it safer to drink? | 1. Yes, always<br>2. Yes, sometime<br>3. No<br>4. Don't know |
| 329 | What do you usually do to make the water safer to drink?<br>*Record all mentioned.* | 1. Boil<br>2. Add bleach/chlorine<br>3. Strain through a cloth<br>4. Use water filter (ceramic/sand/composite/etc.)<br>5. Solar disinfection<br>6. Let it stand and settle<br>7. Other (specify) |
| 330 | What kind of toilet facility do members of your household usually use? | 1. Flush to piped sewer system (not shared with other households)<br>2. Flush to septic tank (not shared with other households)<br>3. No facility/bush/field<br>4. Other type of toilet (specify) |
