## Supplementary material for "Healthcare utilization and expenditure among people with type 2 diabetes and/or hypertension in Cambodia: results from a cross-sectional survey": Annexure 3

### Annexure 3. Individual Survey Questionnaire

**INTRO:**
Hello, my name is ____and I am from the National Institute of Public Health. You has been randomly selected to participate in this study [on the scaling up of diabetes and hypertension in Cambodia] based on information from your household. The information you give will be kept confidential and no personal details will appear in any record. This interview will take approximately 60 minutes. You do not have to answer any question you don't want to and you can stop the interview at any time. There will also be measurements of your blood pressure, weight and height, waist and hip circumferences during and after the interview, and for tomorrow early morning we would like to test your fasting blood glucose [HbA1c and creatinine level for known and suspected diabetes cases]. We very appreciate your participation and information.

| **ELIGIBILITY & RECRUITMENT CHECK**<br>(Instructions: To be filled in by data collectors before participants sign informed consent) | | | | |
|---|---|---|---|---|
| **Q.N** | **Description & questions** | **RESPONSE** | | **Ref:** |
| Q1 | Has the individual been a usual member of the household and stayed in the household the night before the interview or had not been absent for more than 6 months? | 0 = No | 1 = Yes | CDHS |
| Q2 | Is the individual 40 or more than 40 of age?<br><br>*IF POSSIBLE, CHECK ID CARD* | 0 = No | 1 = Yes | WHO PEN SOP |
| | If NO, DO NOT continue. | | | |
| Q3 | Is the individual physically and mentally capable to answer the questions? | 0 = No | 1 = Yes | |
| | If NO, DO NOT continue | | | |
| Q4 | Are you willing to participate in the study? | 0 = No | 1 = Yes | |
| | If NO, DO NOT continue<br>If YES, CONSENT obtained | | | |
| **Q5. Consent obtained** | | 0 = No | 1 = Yes | |

| **SECTION 1: SOCIO-DEMOGRAPHIC INFORMATION** | | | | | |
|---|---|---|---|---|---|
| **Q.N** | **Description & questions** | **RESPONSE** | **Type of Variable** | **Field** | **Ref:** |
| Q6 | How old are you?<br>-*Record in years as stated by the participant*<br>-*Record 99 if don't know* | ____________Years | Quantitative discrete | Age | WHO STEPS |
| Q7 | Sex of participant<br>*Record sex of the participant as observed* | 1 = Male<br>2 = Female | Categorical binary | Sex | WHO STEPS |
| Q8 | What is your marital status? | 1=Married or living together<br>2=Divorced or separated<br>3=Widowed | Categorical nominal | Marital Status | CDHS |

| | | | | | |
|---|---|---|---|---|---|
| | -*Record 88 if refuse to answer* | 4=Never married and never lived together | | | |
| Q9 | What is your highest educational level?<br><br>*Record 99 if don't know* | 1=No formal schooling<br>2=Less than primary school<br>3=Primary school<br>4=Secondary school<br>5=High school<br>6=College/University<br>7=Post graduate degree | Categorical ordinal | Educational level | WHO STEPS |
| Q10 | What is your ethnic group? | 1 = Khmer<br>2 = Vietnamese<br>3 = Chinese<br>4 = Cham (Muslim)<br>5 = Other | Categorical nominal | Ethnicity | WHO STEPS |
| Q11 | Which of the following best describes your main work status within the past 12 months?<br><br>*Record 88 if refuse to answer* | 1=Household tasks<br>2=Civil servant<br>3=Employee of private company/NGO<br>4=Self-employed farmer<br>5=Large-scale farmer with employees<br>6=Self-employed in small business<br>7=Running a big business with employees<br>8=Casual worker<br>9=Working abroad<br>10=At school (pupil/student)<br>11=Unemployed or not eligible<br>12=Retired | Categorical nominal | Occupation | CDHS |
| Q12 | Taking the past year, can you give an estimate of your annual income if I read some options to you?<br><br>*Record 88 if refuse to answer* | 1 = no earnings<br>2 = less than or 250 USD<br>3 = more than 251- 1500 USD<br>4 = more than 1501 – 3500 USD<br>5 = more than 3501 USD | Categorical ordinal | Individual income | WHO STEPS |

### SECTION 2: HEALTH STATUS AND QUALITY OF LIFE

| Q.N | Description and Questions | Response | Type of Variable | Field | Ref: |
|---|---|---|---|---|---|

| | | | | | |
|---|---|---|---|---|---|
| Q13 | At this point of time in your life, how would you describe:<br><br>Q 13.1. Your home situation [ ____ ]<br>Q 13.2. Your family relationships [ ____ ]<br>Q 13.3. Your finances [ ____ ]<br>Q 13.4. Your work situation [ ____ ]<br><br>*-Record 99 if don't know and 88 if refuse* | 1 = Excellent<br>2 = Very good<br>3 = Good<br>4 = Fair<br>5 = Poor | Categorical Ordinal | QoLife | GACD book |
| Q14 | How good or bad is your health today?<br><br>*-The scale is numbered from 0 to 100.*<br>*-100 means the best health you can imagine.*<br>*-0 means the worst health you can imagine.*<br>*-Please mark an X on the scale to indicate how your health is today.*<br><br>*-Record 99 if don't know and 88 if refuse* | 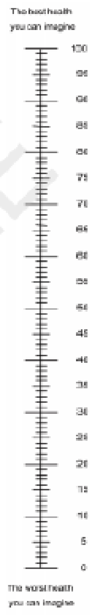 | Quantitative discrete | Gen. Health | EuroQoL |
| Q15 | Have you ever been told by a doctor or other health worker that you have hypertension?<br><br>*-Record 99 if don't know/unsure* | 0 = No<br>1 = Yes | Categorical binary | HT_self-report diagnosis | WHO STEPS |
| Q16 | Have you ever been told by a doctor or other health worker that you have diabetes?<br><br>*-Record 99 if don't know/unsure* | 0 = No<br>1 = Yes | Categorical binary | DM_self-report diagnosis | WHO STEPS |

| | | | | | |
|---|---|---|---|---|---|
| Q17 | Have you ever been told by a doctor or other health worker that you have heart problems?<br><br>*-Record 99 if don't know/unsure* | 0 = No<br>1 = Yes | Categorical binary | Co-morbidity (CVD) | WHO STEPS |
| Q18 | Have you ever been told by a doctor or other health worker that you have symptoms suggestive of a stroke?<br><br>*-Record 99 if don't know/unsure* | 0 = No<br>1 = Yes | Categorical binary | Co-morbidity (CVD) | WHO STEPS |
| Q19 | Have you ever been told by a doctor or other health worker that you have chronic kidney disease?<br><br>*-Record 99 if don't know/unsure* | 0 = No<br>1 = Yes | Categorical binary | Co-morbidity (CKD) | WHO STEPS |
| Q20 | How many natural teeth do you have?<br><br>*-Record 99 if don't know* | 0 = None<br>1 = 1-9 teeth<br>2 = 10-19 teeth<br>3 = 20 teeth or more | Categorical ordinal | Dental health | WHO Oral Health Questionnaire |
| Q21 | During the past 12 months, did your teeth or mouth cause any pain or comfort?<br><br>*-Record 99 if don't know* | 0 = No<br>1 = Yes | Categorical binary | Dental health | WHO Oral Health Questionnaire |
| Q22 | Over the last 2 weeks, how often have you been bothered by any of the following problems?<br><br>Q 22.1. Little interest or pleasure in doing things [ ____ ]<br>Q 22.2. Feeling down, depressed, or hopeless. [ ____ ]<br>Q 22.3. Trouble falling or staying asleep, or sleeping too much. [ ____ ] | 0 = Not at all<br>1 = Several days<br>2 = More than half the days<br>3 = Nearly everyday | Categorical ordinal | Mental health | PHQ-9 |

| | | |
|---|---|---|
| | Q 22.4. Feeling tired or having little energy. [ ____ ]<br>Q 22.5. Poor appetite or overeating [ ____ ]<br>Q 22.6. Feeling bad about yourself – or that you are a failure or make yourself or down your family [ ____ ]<br>Q 22.7. Trouble concentrating on things, such as reading the newspaper or watching television [ ____ ]<br>Q 22.8. Moving or speaking so slowly that other people could have noticed? Or the oppositebeing so fidgety or restless that you have been moving around a lot more than usual [ ____ ]<br>Q 22.9. Thoughts that you would be better off dead or of hurting yourself in some ways [ ____ ] | |
| Q42 | We would like to confirm that:<br><br>*-This question is for categorizing respondents for the following sections and it is also important to ask respondents to confirm their main conditions in this survey.*<br><br>*-IF the answer is "0", go to Section 3*<br>*-IF the answer is "1", go to Section 3a*<br>*-IF the answer is "2", go to Section 3b*<br>*-IF the answer is "3", go to Section 3c* | 0 = Neither hypertension nor diabetes<br><br>1 = Only hypertension<br><br>2 = Only diabetes<br><br>3 = Both diabetes and hypertension |

| SECTION 3: HEALTH CARE UTILIZATION | | | | | |
|---|---|---|---|---|---|
| **Q.N** | **Description and Questions** | **Response** | **Type of Variable** | **Field** | **Ref:** |
| Q23 | Have you sought medical treatment or advice as an outpatient from anyone in the past 3 months?<br><br>*-If No, go to Question 40.* | 0 = No<br>1 = Yes | Categorical variable | Medical advice | GACD book |
| Q24 | Where did you seek medical advice or treatment for illness in the past 3 months?<br><br>*-More than one answer can be selected.*<br>*-Data collectors can use probes to help respondents determine the types of health facilities in the Response Column.*<br>*-Record 99 if don't know and 88 if refuse* | 1= National hospital (PP)<br>2= Provincial hospital (RH)<br>3= District hospital (RH)<br>4= Health centre<br>5= Health post<br>6= Provincial rehabilitation centre (PRC) or Community-based rehabilitation (CBR)<br>7= Other public; specify:<br>8= Private hospital<br>9= Private clinic<br>10= Private pharmacy<br>11= Home/Office of trained health worker/nurse<br>12= Visit of trained health worker/nurse<br>13= Other private medical; specify:<br>14= Shop selling drugs/market<br>15= Kru Khmer/ Magician | Categorical Nominal | Care provider_3M | CDHS and GACD book |

| | | | | | |
|---|---|---|---|---|---|
| | | 16= Monk/religious leader<br>17= Traditional birth attendant<br>18= Oversee medical service<br>19= Other; specify: | | | |
| From Q25-Q39, it is a set of questions that are asked following choices selected in Q24. If 2 or 3 choices were selected in Q24, Q25-Q39 would appear 2 or 3 times, accordingly. | | | | | |
| Q25 | How many times did you visit the selected place**(s)** in Q24 in the past three months? | ________ times | Quantitative discrete | Number of visits | GACD |
| Q26 | How much in total was spent on the treatment at the selected place(s) in Q24?<br><br>-*Record 99 if don't know and 88 if refuse* | 0 = free/no cost<br>1 = in kind<br>2 = _________Riels | -Categorical nominal<br>-<br>Quantitative Continuous | Cost of treatment | CDHS |
| Q27 | How did you pay for the treatment cost at the selected place(s) in Q24?<br><br>-*Record 99 if don't know and 88 if refuse* | 1= Health Equity Fund<br>2= Voucher<br>3= Fee Exemption<br>4= NGO<br>5= National Social Security Fund<br>6= Community-Based Health Insurance<br>7= Health Insurance through Employer<br>8= Other Privately Purchased Commercial Health Insurance | -Categorical nominal | Payment method | GADC adapted to context specific |

| | | | | | |
|---|---|---|---|---|---|
| | | 9= Wage/income<br>10= Loan/ Ton Tin<br>11= Sale of Assets<br>12= Gift from Relative<br>13= Savings<br>14= Other | | | |
| Q28 | How much in total was spent on transport to go to and return from the selected place(s) in Q24?<br><br>-*Record 99 if don't know and 88 if refuse* | 0 = free/no cost<br>1 = in kind<br>2 = ________Riels | -Categorical nominal<br>-<br>Quantitative Continuous | Cost of transp ort | CDHS |
| Q29 | On average how many hours do you spend to get treatment/advices from the selected place(s) in Q24?<br><br>-*Record 99 if don't know and 88 if refuse* | ________Hours | Quantitative continuous | Time spendi ng | GACD book (Adapted) |
| Q30 | How satisfied are you with the effect of your {treatment/care} at the selected place(s) in Q24?<br><br>-*Record 99 if don't know/unsure* | 0 = Very satisfied<br>1 = Satisfied<br>2 = Neither satisfied nor dissatisfied<br>3 = Dissatisfied<br>4 = Very dissatisfied | Categorical ordinal | Patien t satisfa ction of care servic es | SAPS 2006 |
| Q31 | How satisfied are you with the explanations the {doctor/other health professional} has given you about the results of your {treatment/care} at the selected place(s) in Q24?<br><br>-*Record 99 if don't know/unsure* | 0 = Very satisfied<br>1 = Satisfied<br>2 = Neither satisfied nor dissatisfied<br>3 = Dissatisfied<br>4 = Very dissatisfied | Categorical ordinal | Patien t satisfa ction of care servic es | SAPS 2006 |

| | | | | | |
|---|---|---|---|---|---|
| Q32 | The {doctor/other health professional} at the selected place(s) in Q24 was very careful to check everything when examining you.<br><br>*-Record 99 if don't know/unsure* | 0 = Strongly agree<br>1 = Agree<br>2 = Not sure<br>3 = Disagree<br>4 = Strongly disagree | Categorical ordinal | Patient satisfaction of care services | SAPS 2006 |
| Q33 | At the selected place(s) in Q24, how satisfied were you with the choices you had in decisions affecting your health care?<br><br>*-Record 99 if don't know/unsure* | 0 = Very satisfied<br>1 = Satisfied<br>2 = Neither s atisfied nor dissatisfied<br>3 = Dissatisfied<br>4 = Very dissatisfied | Categorical ordinal | Patient satisfaction of care services | SAPS 2006 |
| Q34 | How much of the time did you feel respected by the {doctor/other health professional} at the selected place(s) in Q24?<br><br>*-Record 99 if don't know/unsure* | 0 = All of the time<br>1 = Most of the time<br>2 = About half the time<br>3 = Some of the time<br>4 = None of the time | Categorical ordinal | Patient satisfaction of care services | SAPS 2006 |
| Q35 | At the selected place(s) in Q24, the time you had with the {doctor/other health professional} was too short.<br><br>*-Record 99 if don't know/unsure* | 0 = Strongly agree<br>1 = Agree<br>2 = Not sure<br>3 = Disagree<br>4 = Strongly disagree | Categorical ordinal | Patient satisfaction of care services | SAPS 2006 |
| Q36 | Are you satisfied with the care you received in the selected place(s) in Q24?<br><br>*-Record 99 if don't know/unsure* | 0 = Very satisfied<br>1 = Satisfied<br>2 = Neither s atisfied nor dissatisfied<br>3 = Dissatisfied<br>4 = Very dissatisfied | Categorical ordinal | Patient satisfaction of care services | SAPS 2006 |

| | | | | | |
|---|---|---|---|---|---|
| Q37 | Did you get your blood pressure measured at the selected place(s) in Q24?<br><br>-*Record 99 if don't know/unsure* | 0 = No<br>1 = Yes | Categorical variable | Access to blood pressure testing | Opinion |
| Q38 | Did you get your blood glucose tested at the selected place(s) in Q24?<br><br>-*Record 99 if don't know/unsure* | 0 = No<br>1 = Yes | Categorical variable | Access to blood glucose testing | Opinion |
| Q39 | Would you recommend the selected place(s) in Q24 to others?<br><br>-*Record 99 if don't know/unsure* | 1 = Not recommended<br>2 = Recommend with reservations<br>3 = Recommend<br>4 = Highly recommend | Categorical ordinal | Patient satisfaction of care services | Opinion |
| Q40 | Have you ever had your blood glucose tested in the last three years?<br><br>-*Record 99 if don't know/unsure* | 0 = No<br>1 = Yes | Categorical binary | Testing DM (CoC) | Opinion |
| Q41 | Have you ever had your blood pressure measured in the last three years?<br><br>-*Record 99 if don't know/unsure* | 0 = No<br>1 = Yes | Categorical binary | Testing HT (CoC) | Opinion |

| SECTION 3a: HEALTH CARE UTILIZATION FOR HYPERTENSION | | | | | |
|---|---|---|---|---|---|
| **Q.N** | **Description and Questions** | **Response** | **Type of Variable** | **Field** | **Ref:** |
| Q43a | How long have you lived with hypertension?<br><br>*-Record 99 if don't know/unsure and 88 if refuse*<br>*-Less than a year is rounded up to one year*<br>*-Standard rounded up formula is applied.* | ________Years | Quantitative concrete | Duration HT | Opinion |
| Q44a | Where were you first diagnosed as having hypertension?<br><br>*-Record 99 if don't know and 88 if refuse* | 1= National hospital (PP)<br>2= Provincial hospital (RH)<br>3= District hospital (RH)<br>4= Health centre<br>5= Health post<br>6= Provincial rehabilitation centre (PRC) or Community-based rehabilitation (CBR)<br>7= Other public; specify:<br>8= Private hospital<br>9= Private clinic<br>10= Private pharmacy<br>11= Home/Office of trained health worker/nurse<br>12= Visit of trained health worker/nurse<br>13= Other private medical; specify:<br>14= Shop selling drugs/market<br>15= Kru Khmer/ Magician | Categorical Nominal | Diagnosis HT (CoC) | Opinion |

| | | | | | |
|---|---|---|---|---|---|
| | | 16= Monk/religious leader<br>17= Traditional birth attendant<br>18= Oversee medical service<br>19= MoPoTsyo<br>20= Other; specify: | | | |
| Q45a | Where did you first seek advice or treatment for hypertension after being diagnosed?<br><br>*-Record 99 if don't know and 88 if refuse* | 1= National hospital (PP)<br>2= Provincial hospital (RH)<br>3= District hospital (RH)<br>4= Health centre<br>5= Health post<br>6= Provincial rehabilitation centre (PRC) or Community-based rehabilitation (CBR)<br>7= Other public; specify:<br>8= Private hospital<br>9= Private clinic<br>10= Private pharmacy<br>11= Home/Office of trained health worker/nurse<br>12= Visit of trained health worker/nurse<br>13= Other private medical; specify:<br>14= Shop selling drugs/market<br>15= Kru Khmer/ Magician<br>16= Monk/religious leader<br>17= Traditional birth attendant | Categorical Nominal | Link to care HT (CoC) | CDHS (Adapted for disease specific) |

| | | | | | |
|---|---|---|---|---|---|
| | | 18= Oversee medical service<br>19= MoPoTsyo<br>20= Other; specify: | | | |
| Q46a | Did you go to other places for follow up treatment/care for your hypertensive conditions?<br><br>*-Record 88 if refuse*<br>*-If NO, please skip Q47a* | 0 = No<br>1 = Yes | Categorical binary | Trust HT | CDHS |
| Q47a | If yes to Q46a, where else did you go to get follow up treatment/care for your hypertensive conditions? | 1= National hospital (PP)<br>2= Provincial hospital (RH)<br>3= District hospital (RH)<br>4= Health centre<br>5= Health post<br>6= Provincial rehabilitation centre (PRC) or Community-based rehabilitation (CBR)<br>7= Other public; specify:<br>8= Private hospital<br>9= Private clinic<br>10= Private pharmacy<br>11= Home/Office of trained health worker/nurse<br>12= Visit of trained health worker/nurse<br>13= Other private medical; specify:<br>14= Shop selling drugs/market | Categorical Nominal | Link to care HT (CoC) | CDHS (Adapted for disease specific) |

| | | | | | |
|---|---|---|---|---|---|
| | | 15= Kru Khmer/ Magician<br>16= Monk/religious leader<br>17= Traditional birth attendant<br>18= Oversee medical service<br>19= MoPoTsyo<br>20= Other; specify: | | | |
| Q48a | Did you get treatment/care for your hypertensive conditions in the past **12 months?**<br><br>*-Record 88 if refuse*<br>*-If NO, please skip Q49a-64a* | 0 = No<br>1 = Yes | Categorical binary | Retain in care HT (CoC) | CDHS (Adapted for disease specific) |
| Q49a | Are you currently receiving any of the following treatment/advices for your hypertensive conditions prescribed by a doctor or other health care worker?<br><br>Q 49.1a. Drugs (medication) that you have taken in the past two weeks [ ____ ]<br>Q 49.2a. Advice to reduce salt intake [ ___ ]<br>Q 49.3a. Advice or treatment to lose weight [ ____ ]<br>Q 49.4a. Advice or treatment to stop smoking [ ____ ]<br>Q 49.5a. Advice to start or do more physical exercise [ ____ ]<br><br>*-Record 99 if don't know and 88 if refuse* | 0 = No 1= Yes | Categorical ordinal | In treatment for HT (CoC) | STEPS Survey |
| Q50a | Have you had your blood cholesterol measured in the past 12 months?<br><br>*-Record 99 if don't know and 88 if refuse* | 0 = No 1= Yes | Categorical ordinal | In treatment for HT (CoC) | Veerle's suggestion |

| | | | | | |
|---|---|---|---|---|---|
| Q51a | Where did you seek medical advice or treatment for your hypertensive condition in **the past 3 months**?<br><br>-***More than one answer can be selected***.<br>-*Data collectors can use probes to help respondents determine the types of health facilities in the Response Column.*<br>-*Record 99 if don't know and 88 if refuse*<br>-*If (21=no wehere), go to Q65a* | 1= National hospital (PP)<br>2= Provincial hospital (RH)<br>3= District hospital (RH)<br>4= Health centre<br>5= Health post<br>6= Provincial rehabilitation centre (PRC) or Community-based rehabilitation (CBR)<br>7= Other public; specify:<br>8= Private hospital<br>9= Private clinic<br>10= Private pharmacy<br>11= Home/Office of trained health worker/nurse<br>12= Visit of trained health worker/nurse<br>13= Other private medical; specify:<br>14= Shop selling drugs/market<br>15= Kru Khmer/ Magician<br>16= Monk/religious leader<br>17= Traditional birth attendant<br>18= Oversee medical service<br>19= MoPoTsyo<br>20= Other; specify: | Categorical Nominal | Care provider_3M | CDHS and GACD book |

| | | | | | |
|---|---|---|---|---|---|
| | | 21= No where | | | |
| From Q52a-Q64a, it is a set of questions that are asked following choices selected in Q51a. If 2 or 3 choices were selected in Q51a, Q52a-Q64a would appear 2 or 3 times, accordingly. | | | | | |
| Q52a | How many times did you visit the selected place**(s)** in Q51a in the past three months? | ________ times | Quantitative discrete | Numb er of visits | GACD |
| Q53a | How much in total was spent on the treatment at the selected place(s) in Q51a?<br><br>-*Record 99 if don't know and 88 if refuse* | 0 = free/no cost<br>1 = in kind<br>2 = _________Riels | -Categorical nominal<br>-Quantitative Continuous | Cost of treatm ent | CDHS |
| Q54a | How did you pay for the treatment cost at the selected place(s) in Q51a?<br><br>-*Record 99 if don't know and 88 if refuse* | 1= Health Equity Fund<br>2= Voucher<br>3= Fee Exemption<br>4= NGO<br>5= National Social Security Fund<br>6= Community-Based Health Insurance<br>7= Health Insurance through Employer<br>8= Other Privately Purchased Commercial Health Insurance | -Categorical nominal | Paym ent metho d | GADC adapted to context specific |

| | | | | | |
|---|---|---|---|---|---|
| | | 9= Wage/income<br>10= Loan/ Ton Tin<br>11= Sale of Assets<br>12= Gift from Relative<br>13= Savings<br>14= Other | | | |
| Q55a | How much in total was spent on transport to go to and return from the selected place(s) in Q51a?<br><br>-*Record 99 if don't know and 88 if refuse* | 0 = free/no cost<br>1 = in kind<br>2 = _________Riels | -Categorical nominal<br>-<br>Quantitative Continuous | Cost of transp ort | CDHS |
| Q56a | On average how many hours do you spend to get treatment/advices from the selected place(s) in Q51a?<br><br>-*Record 99 if don't know and 88 if refuse* | _________Hours | Quantitative continuous | Time spendi ng | GACD book (Adapted) |
| Q57a | How satisfied are you with the effect of your {treatment/care} at the selected place(s) in Q51a?<br><br>-*Record 99 if don't know/unsure* | 0 = Very satisfied<br>1 = Satisfied<br>2 = Neither satisfied nor dissatisfied<br>3 = Dissatisfied<br>4 = Very dissatisfied | Categorical ordinal | Patien t satisfa ction of care servic es | SAPS 2006 |
| Q58a | How satisfied are you with the explanations the {doctor/other health professional} has given you about the results of your {treatment/care} at the selected place(s) in Q51a?<br><br>-*Record 99 if don't know/unsure* | 0 = Very satisfied<br>1 = Satisfied<br>2 = Neither satisfied nor dissatisfied<br>3 = Dissatisfied<br>4 = Very d issatisfied | Categorical ordinal | Patien t satisfa ction of care servic es | SAPS 2006 |

| | | | | | |
|---|---|---|---|---|---|
| Q59a | The {doctor/other health professional} at the selected place(s) in Q51a was very careful to check everything when examining you.<br><br>*-Record 99 if don't know/unsure* | 0 = Strongly agree<br>1 = Agree<br>2 = Not sure<br>3 = Disagree<br>4 = Strongly disagree | Categorical ordinal | Patient satisfaction of care services | SAPS 2006 |
| Q60a | At the selected place(s) in Q51a, how satisfied were you with the choices you had in decisions affecting your health care?<br><br>*-Record 99 if don't know/unsure* | 0 = Very satisfied<br>1 = Satisfied<br>2 = Neither satisfied nor dissatisfied<br>3 = Dissatisfied<br>4 = Very dissatisfied | Categorical ordinal | Patient satisfaction of care services | SAPS 2006 |
| Q61a | How much of the time did you feel respected by the {doctor/other health professional} at the selected place(s) in Q51a?<br><br>*-Record 99 if don't know/unsure* | 0 = All of the time<br>1 = Most of the time<br>2 = About half the time<br>3 = Some of the time<br>4 = None of the time | Categorical ordinal | Patient satisfaction of care services | SAPS 2006 |
| Q62a | At the selected place(s) in Q51a, the time you had with the {doctor/other health professional} was too short.<br><br>*-Record 99 if don't know/unsure* | 0 = Strongly agree<br>1 = Agree<br>2 = Not sure<br>3 = Disagree<br>4 = Strongly disagree | Categorical ordinal | Patient satisfaction of care services | SAPS 2006 |
| Q63a | Are you satisfied with the care you received in the selected place(s) in Q51a?<br><br>*-Record 99 if don't know/unsure* | 0 = Very satisfied<br>1 = Satisfied<br>2 = Neither satisfied nor dissatisfied<br>3 = Dissatisfied<br>4 = Very dissatisfied | Categorical ordinal | Patient satisfaction of care services | SAPS 2006 |

| | | | | | |
|---|---|---|---|---|---|
| Q64a | Would you recommend the selected place(s) in Q51a to others?<br><br>-*Record 99 if don't know/unsure* | 1 = Not recommend<br>2 = Recommend with reservations<br>3 = Recommend<br>4 = Highly recommend | Categorical ordinal | Patient satisfaction of care services | Opinion |
| Q65a | Have you ever been told by a doctor that you have eyes problems?<br><br>-*Record 99 if don't know/unsure* | 0 = No<br>1 = Yes | Quantitative discrete | Complication-HT | Cambodia national guideline |
| Q66a | Have you ever been told by a doctor that you have kidney problems?<br><br>-*Record 99 if don't know/unsure* | 0 = No<br>1 = Yes | Quantitative discrete | Complication-HT | Cambodia national guideline |
| Q67a | Have you ever been told by a doctor that you have lost the sensation of your peripheral membrane, such as foot, hand, arm?<br><br>-*Record 99 if don't know/unsure* | 0 = No<br>1 = Yes | Quantitative discrete | Complication-HT | Cambodia national guideline |
| | In **the past 3 months**, besides seeking medical advice or treatment for your hypertensive condition, have you sought medical treatment or advice for other illnesses or conditions?<br><br>If No, go to Section 4. | 0 = No<br>1 = Yes | | | |

| | | | | | |
|---|---|---|---|---|---|
| Q24 | Where did you seek medical advice or treatment for that illness in the past 3 months?<br><br>*-More than one answer can be selected.*<br>*-Data collectors can use probes to help respondents determine the types of health facilities in the Response Column.*<br>*-Record 99 if don't know and 88 if refuse* | 1= National hospital (PP)<br>2= Provincial hospital (RH)<br>3= District hospital (RH)<br>4= Health centre<br>5= Health post<br>6= Provincial rehabilitation centre (PRC) or Community-based rehabilitation (CBR)<br>7= Other public; specify:<br>8= Private hospital<br>9= Private clinic<br>10= Private pharmacy<br>11= Home/Office of trained health worker/nurse<br>12= Visit of trained health worker/nurse<br>13= Other private medical; specify:<br>14= Shop selling drugs/market<br>15= Kru Khmer/ Magician<br>16= Monk/religious leader<br>17= Traditional birth attendant<br>18= Oversee medical service<br>19= Other; specify: | Categorical Nominal | Care provider_3M | CDHS and GACD book |
| From Q25-Q39, it is a set of questions that are asked following choices selected in Q24. If 2 or 3 choices were selected in Q24, Q25-Q39 would appear 2 or 3 times, accordingly. | | | | | |

| | | | | | |
|---|---|---|---|---|---|
| Q25 | How many times did you visit the selected place**(s)** in Q24 in the past three months? | _______ times | Quantitative discrete | Number of visits | GACD |
| Q26 | How much in total was spent on the treatment at the selected place(s) in Q24?<br><br>-*Record 99 if don't know and 88 if refuse* | 0 = free/no cost<br>1 = in kind<br>2 = ________Riels | -Categorical nominal<br>-Quantitative Continuous | Cost of treatment | CDHS |
| Q27 | How did you pay for the treatment cost at the selected place(s) in Q24?<br><br>-*Record 99 if don't know and 88 if refuse* | 1= Health Equity Fund<br>2= Voucher<br>3= Fee Exemption<br>4= NGO<br>5= National Social Security Fund<br>6= Community-Based Health Insurance<br>7= Health Insurance through Employer<br>8= Other Privately Purchased Commercial Health Insurance<br>9= Wage/income<br>10= Loan/ Ton Tin<br>11= Sale of Assets<br>12= Gift from Relative<br>13= Savings<br>14= Other | -Categorical nominal | Payment method | GADC adapted to context specific |

| | | | | | |
|---|---|---|---|---|---|
| Q28 | How much in total was spent on transport to go to and return from the selected place(s) in Q24?<br><br>*-Record 99 if don't know and 88 if refuse* | 0 = free/no cost<br>1 = in kind<br>2 = ________Riels | -Categorical nominal<br>-Quantitative Continuous | Cost of transp ort | CDHS |
| Q29 | On average how many hours do you spend to get treatment/advices from the selected place(s) in Q24?<br><br>*-Record 99 if don't know and 88 if refuse* | ________Hours | Quantitative continuous | Time spendi ng | GACD book (Adapted) |
| Q30 | How satisfied are you with the effect of your {treatment/care} at the selected place(s) in Q24?<br><br>*-Record 99 if don't know/unsure* | 0 = Very satisfied<br>1 = Satisfied<br>2 = Neither satisfied nor dissatisfied<br>3 = Dissatisfied<br>4 = Very dissatisfied | Categorical ordinal | Patien t satisfa ction of care servic es | SAPS 2006 |
| Q31 | How satisfied are you with the explanations the {doctor/other health professional} has given you about the results of your {treatment/care} at the selected place(s) in Q24?<br><br>*-Record 99 if don't know/unsure* | 0 = Very satisfied<br>1 = Satisfied<br>2 = Neither satisfied nor dissatisfied<br>3 = Dissatisfied<br>4 = Very dissatisfied | Categorical ordinal | Patien t satisfa ction of care servic es | SAPS 2006 |
| Q32 | The {doctor/other health professional} at the selected place(s) in Q24 was very careful to check everything when examining you.<br><br>*-Record 99 if don't know/unsure* | 0 = Strongly agree<br>1 = Agree<br>2 = Not sure<br>3 = Disagree<br>4 = Strongly disagree | Categorical ordinal | Patien t satisfa ction of care servic es | SAPS 2006 |

| | | | | | |
|---|---|---|---|---|---|
| Q33 | At the selected place(s) in Q24, how satisfied were you with the choices you had in decisions affecting your health care?<br><br>*-Record 99 if don't know/unsure* | 0 = Very satisfied<br>1 = Satisfied<br>2 = Neither s atisfied nor dissatisfied<br>3 = Dissatisfied<br>4 = Very dissatisfied | Categorical ordinal | Patien t satisfa ction of care servic es | SAPS 2006 |
| Q34 | How much of the time did you feel respected by the {doctor/other health professional} at the selected place(s) in Q24?<br><br>*-Record 99 if don't know/unsure* | 0 = All of the time<br>1 = Most of the time<br>2 = About half the time<br>3 = Some of the time<br>4 = None of the time | Categorical ordinal | Patien t satisfa ction of care servic es | SAPS 2006 |
| Q35 | At the selected place(s) in Q24, the time you had with the {doctor/other health professional} was too short.<br><br>*-Record 99 if don't know/unsure* | 0 = Strongly agree<br>1 = Agree<br>2 = Not sure<br>3 = Disagree<br>4 = Strongly disagree | Categorical ordinal | Patien t satisfa ction of care servic es | SAPS 2006 |
| Q36 | Are you satisfied with the care you received in the selected place(s) in Q24?<br><br>*-Record 99 if don't know/unsure* | 0 = Very satisfied<br>1 = Satisfied<br>2 = Neither s atisfied nor dissatisfied<br>3 = Dissatisfied<br>4 = Very dissatisfied | Categorical ordinal | Patien t satisfa ction of care servic es | SAPS 2006 |
| Q37 | Did you get your blood pressure measured at the selected place(s) in Q24?<br><br>*-Record 99 if don't know/unsure* | 0 = No<br>1 = Yes | Categorical variable | Acces s to blood pressu re testing | Opinion |

| | | | | | |
|---|---|---|---|---|---|
| Q38 | Did you get your blood glucose tested at the selected place(s) in Q24?<br><br>*-Record 99 if don't know/unsure* | 0 = No<br>1 = Yes | Categorical variable | Access to blood glucose testing | Opinion |
| Q39 | Would you recommend the selected place(s) in Q24 to others?<br><br>*-Record 99 if don't know/unsure* | 1 = Not recommended<br>2 = Recommend with reservations<br>3 = Recommend<br>4 = Highly recommend | Categorical ordinal | Patient satisfaction of care services | Opinion |

| SECTION 3b: HEALTH CARE UTILIZATION FOR DIABETES | | | | | |
|---|---|---|---|---|---|
| **Q.N** | **Description and Questions** | **Response** | **Type of Variable** | **Field** | **Ref:** |
| Q43b | How long have you lived with diabetes?<br><br>*-Record 99 if don't know/unsure and 88 if refuse*<br>*-Less than a year is rounded up to one year*<br>*-Standard rounded up formula is applied.* | ________Years | Quantitative concrete | Duration DM | Opinion |
| Q44b | Where were you first diagnosed as having diabetes?<br><br>*-Record 99 if don't know and 88 if refuse* | 1= National hospital (PP)<br>2= Provincial hospital (RH)<br>3= District hospital (RH)<br>4= Health centre<br>5= Health post<br>6= Provincial rehabilitation centre (PRC) or Community-based rehabilitation (CBR)<br>7= Other public; specify:<br>8= Private hospital<br>9= Private clinic<br>10= Private pharmacy<br>11= Home/Office of trained health worker/nurse<br>12= Visit of trained health worker/nurse<br>13= Other private medical; specify:<br>14= Shop selling drugs/market<br>15= Kru Khmer/ Magician | Categorical Nominal | Diagnosis DM (CoC) | Opinion |

| | | | | | |
|---|---|---|---|---|---|
| | | 16= Monk/religious leader<br>17= Traditional birth attendant<br>18= Oversee medical service<br>19= MoPoTsyo<br>20= Other; specify: | | | |
| Q45b | Where did you first seek advice or treatment for diabetes after being diagnosed?<br><br>-*Record 99 if don't know and 88 if refuse* | 1= National hospital (PP)<br>2= Provincial hospital (RH)<br>3= District hospital (RH)<br>4= Health centre<br>5= Health post<br>6= Provincial rehabilitation centre (PRC) or Community-based rehabilitation (CBR)<br>7= Other public; specify:<br>8= Private hospital<br>9= Private clinic<br>10= Private pharmacy<br>11= Home/Office of trained health worker/nurse<br>12= Visit of trained health worker/nurse<br>13= Other private medical; specify:<br>14= Shop selling drugs/market<br>15= Kru Khmer/ Magician<br>16= Monk/religious leader<br>17= Traditional birth attendant | Categorical Nominal | Link to care DM (CoC) | CDHS (Adapted for disease specific) |

| | | | | | |
|---|---|---|---|---|---|
| | | 18= Oversee medical service<br>19= MoPoTsyo<br>20= Other; specify: | | | |
| Q46b | Did you go to other places for follow up treatment/care for your diabetes conditions?<br><br>*-Record 88 if refuse*<br>*-If NO, please skip Q51b* | 0 = No<br>1 = Yes | Categorical binary | Trust DM | CDHS |
| Q47b | If yes to Q46b, where else did you go to get follow up treatment/care for your diabetes conditions? | 1= National hospital (PP)<br>2= Provincial hospital (RH)<br>3= District hospital (RH)<br>4= Health centre<br>5= Health post<br>6= Provincial rehabilitation centre (PRC) or Community-based rehabilitation (CBR)<br>7= Other public; specify:<br>8= Private hospital<br>9= Private clinic<br>10= Private pharmacy<br>11= Home/Office of trained health worker/nurse<br>12= Visit of trained health worker/nurse<br>13= Other private medical; specify:<br>14= Shop selling drugs/market | Categorical Nominal | Link to care DM (CoC) | CDHS (Adapted for disease specific) |

| | | | | | |
|---|---|---|---|---|---|
| | | 15= Kru Khmer/ Magician<br>16= Monk/religious leader<br>17= Traditional birth attendant<br>18= Oversee medical service<br>19= MoPoTsyo<br>20= Other; specify: | | | |
| Q48b | Did you get treatment/care for your diabetes conditions in the past 12 months?<br><br>*-Record 88 if refuse*<br>*-If NO, please skip Q49b-65b* | 0 = No<br>1 = Yes | Categorical binary | Retain in care DM (CoC) | CDHS (Adapted for disease specific) |
| Q49b | Are you currently receiving any of the following treatment/advices for your diabetes conditions prescribed by a doctor or other health care worker?<br><br>Q. 49b.1. Insulin [ ____ ]<br>Q. 49b.2. Drugs (medication) that you have taken in the past two weeks [ ____ ]<br>Q. 49b.3. Special prescribed diet [ ____ ]<br>Q. 49b.4. Advice or treatment to lose weight [ ____ ]<br>Q. 49b.5. Advice or treatment to stop smoking<br>Q. 49b.6. Advice to start or do more physical exercise | 0 = No 1= Yes | Categorical ordinal | In treatm ent for DM (CoC) | STEPS Survey |
| Q50b | Have you had your blood glucose measured in the past 12 months?<br><br>*-Record 99 if don't know and 88 if refuse* | 0 = No 1= Yes | Categorical ordinal | In treatm ent for DM (CoC) | Veerle's suggestion |

| | | | | | |
|---|---|---|---|---|---|
| Q51b | Have you had your HbA1c tested in the past 12 months?<br><br>*-Record 99 if don't know and 88 if refuse* | 0 = No 1= Yes | Categorical ordinal | In treatment for DM (CoC) | Veerle's suggestion |
| Q52b | Where did you seek medical advice or treatment for illness in the **past 3 months**?<br><br>***-More than one answer can be selected.***<br>*-Data collectors can use probes to help respondents determine the types of health facilities in the Response Column.*<br>*-Record 99 if don't know and 88 if refuse*<br>*-If 21=Nowhere, go to Q66b.* | 1= National hospital (PP)<br>2= Provincial hospital (RH)<br>3= District hospital (RH)<br>4= Health centre<br>5= Health post<br>6= Provincial rehabilitation centre (PRC) or Community-based rehabilitation (CBR)<br>7= Other public; specify:<br>8= Private hospital<br>9= Private clinic<br>10= Private pharmacy<br>11= Home/Office of trained health worker/nurse<br>12= Visit of trained health worker/nurse<br>13= Other private medical; specify:<br>14= Shop selling drugs/market<br>15= Kru Khmer/ Magician<br>16= Monk/religious leader<br>17= Traditional birth attendant<br>18= Oversee medical service | Categorical Nominal | Care provider_3M | CDHS and GACD book |

| | | | | | |
|---|---|---|---|---|---|
| | | 19= MoPoTsyo<br>20= Other; specify:<br>21= No where | | | |
| From Q53b-Q65b, it is a set of questions that are asked following choices selected in Q52b. If 2 or 3 choices were selected in Q52b, Q53b-Q65b would appear 2 or 3 times, accordingly. | | | | | |
| Q53b | How many times did you visit the selected place**(s)** in Q52b in the past three months? | ________ times | Quantitative discrete | Number of visits | GACD |
| Q54b | How much in total was spent on the treatment at the selected place(s) in Q52b?<br><br>-*Record 99 if don't know and 88 if refuse* | 0 = free/no cost<br>1 = in kind<br>2 = _________Riels<br>OR<br>_________ USD | -Categorical nominal<br>-<br>Quantitative Continuous | Cost of treatment | CDHS |
| Q55b | How did you pay for the treatment cost at the selected place(s) in Q52b?<br><br>-*Record 99 if don't know and 88 if refuse* | 1= Health Equity Fund<br>2= Voucher<br>3= Fee Exemption<br>4= NGO<br>5= National Social Security Fund<br>6= Community-Based Health Insurance<br>7= Health Insurance through Employer<br>8= Other Privately | -Categorical nominal | Payment method | GADC adapted to context specific |

| | | | | | |
|---|---|---|---|---|---|
| | | Purchased Commercial Health Insurance<br>9= Wage/income<br>10= Loan/ Ton Tin<br>11= Sale of Assets<br>12= Gift from Relative<br>13= Savings<br>14= Other | | | |
| Q56b | How much in total was spent on transport to go to and return from the selected place(s) in Q52b?<br><br>-*Record 99 if don't know and 88 if refuse* | 0 = free/no cost<br>1 = in kind<br>2 = _________Riels<br>OR<br>_________ USD | -Categorical nominal<br>-<br>Quantitative Continuous | Cost of transp ort | CDHS |
| Q57b | On average how many hours do you spend to get treatment/advices from the selected place(s) in Q52b?<br><br>-*Record 99 if don't know and 88 if refuse* | _________Hours | Quantitative continuous | Time spendi ng | GACD book (Adapted) |
| Q58b | How satisfied are you with the effect of your {treatment/care} at the selected place(s) in Q52b?<br><br>-*Record 99 if don't know/unsure* | 0 = Very satisfied<br>1 = Satisfied<br>2 = Neither satisfied nor dissatisfied<br>3 = Dissatisfied<br>4 = Very dissatisfied | Categorical ordinal | Patien t satisfa ction of care servic e | SAPS 2006 |
| Q59b | How satisfied are you with the explanations the {doctor/other health professional} has given you about the results of your {treatment/care} at the selected place(s) in Q52b?<br><br>-*Record 99 if don't know/unsure* | 0 = Very satisfied<br>1 = Satisfied<br>2 = Neither satisfied nor dissatisfied<br>3 = Dissatisfied<br>4 = Very d issatisfied | Categorical ordinal | Patien t satisfa ction of care servic e | SAPS 2006 |

| | | | | | |
|---|---|---|---|---|---|
| Q60b | The {doctor/other health professional} at the selected place(s) in Q52b was very careful to check everything when examining you.<br><br>*-Record 99 if don't know/unsure* | 0 = Strongly agree<br>1 = Agree<br>2 = Not sure<br>3 = Disagree<br>4 = Strongly disagree | Categorical ordinal | Patient satisfaction of care service | SAPS 2006 |
| Q61b | At the selected place(s) in Q52b, how satisfied were you with the choices you had in decisions affecting your health care?<br><br>*-Record 99 if don't know/unsure* | 0 = Very satisfied<br>1 = Satisfied<br>2 = Neither satisfied nor dissatisfied<br>3 = Dissatisfied<br>4 = Very dissatisfied | Categorical ordinal | Patient satisfaction of care service | SAPS 2006 |
| Q62b | How much of the time did you feel respected by the {doctor/other health professional} at the selected place(s) in Q52b?<br><br>*-Record 99 if don't know/unsure* | 0 = All of the time<br>1 = Most of the time<br>2 = About half the time<br>3 = Some of the time<br>4 = None of the time | Categorical ordinal | Patient satisfaction of care service | SAPS 2006 |
| Q63b | At the selected place(s) in Q52b, the time you had with the {doctor/other health professional} was too short.<br><br>*-Record 99 if don't know/unsure* | 0 = Strongly agree<br>1 = Agree<br>2 = Not sure<br>3 = Disagree<br>4 = Strongly disagree | Categorical ordinal | Patient satisfaction of care service | SAPS 2006 |
| Q64b | Are you satisfied with the care you received in the selected place(s) in Q52b?<br><br>*-Record 99 if don't know/unsure* | 0 = Very satisfied<br>1 = Satisfied<br>2 = Neither satisfied nor dissatisfied<br>3 = Dissatisfied<br>4 = Very dissatisfied | Categorical ordinal | Patient satisfaction of care service | SAPS 2006 |

| | | | | | |
|---|---|---|---|---|---|
| Q65b | Would you recommend the selected place(s) in Q52b to others?<br><br>*-Record 99 if don't know/unsure* | 1 = Not recommended<br>2 = Recommend with reservations<br>3 = Recommend<br>4 = Highly recommend | Categorical ordinal | Patient satisfaction of care services | Opinion |
| Q66b | Have you ever been told by a doctor that you have eyes problems?<br><br>*-Record 99 if don't know/unsure* | 0 = No<br>1 = Yes | Quantitative discrete | Complication-DM | Cambodia national guideline |
| Q67b | Have you ever been told by a doctor that you have kidney problems?<br><br>*-Record 99 if don't know/unsure* | 0 = No<br>1 = Yes | Quantitative discrete | Complication-DM | Cambodia national guideline |
| Q68b | Have you ever been told by a doctor that you have lost the sensation of your peripheral membrane, such as foot, hand, arm?<br><br>*-Record 99 if don't know/unsure* | 0 = No<br>1 = Yes | Quantitative discrete | Complication-DM | Cambodia national guideline |
| | In **the past 3 months**, besides seeking medical advice or treatment for your diabetes condition, have you sought medical treatment or advice for other illnesses or conditions?<br><br>If No, go to Section 4. | 0 = No<br>1 = Yes | | | |

| | | | | | |
|---|---|---|---|---|---|
| Q24 | Where did you seek medical advice or treatment for that illness in the past 3 months?<br><br>*-More than one answer can be selected.*<br>*-Data collectors can use probes to help respondents determine the types of health facilities in the Response Column.*<br>*-Record 99 if don't know and 88 if refuse* | 1= National hospital (PP)<br>2= Provincial hospital (RH)<br>3= District hospital (RH)<br>4= Health centre<br>5= Health post<br>6= Provincial rehabilitation centre (PRC) or Community-based rehabilitation (CBR)<br>7= Other public; specify:<br>8= Private hospital<br>9= Private clinic<br>10= Private pharmacy<br>11= Home/Office of trained health worker/nurse<br>12= Visit of trained health worker/nurse<br>13= Other private medical; specify:<br>14= Shop selling drugs/market<br>15= Kru Khmer/ Magician<br>16= Monk/religious leader<br>17= Traditional birth attendant<br>18= Oversee medical service<br>19= Other; specify: | Categorical Nominal | Care provider_3M | CDHS and GACD book |
| From Q25-Q39, it is a set of questions that are asked following choices selected in Q24. If 2 or 3 choices were selected in Q24, Q25-Q39 would appear 2 or 3 times, accordingly. | | | | | |

| | | | | | |
|---|---|---|---|---|---|
| Q25 | How many times did you visit the selected place**(s)** in Q24 in the past three months? | ________ times | Quantitative discrete | Numb er of visits | GACD |
| Q26 | How much in total was spent on the treatment at the selected place(s) in Q24?<br><br>*-Record 99 if don't know and 88 if refuse* | 0 = free/no cost<br>1 = in kind<br>2 = _________Riels | -Categorical nominal<br>-<br>Quantitative Continuous | Cost of treatm ent | CDHS |
| Q27 | How did you pay for the treatment cost at the selected place(s) in Q24?<br><br>*-Record 99 if don't know and 88 if refuse* | 1= Health Equity Fund<br>2= Voucher<br>3= Fee Exemption<br>4= NGO<br>5= National Social Security Fund<br>6= Community-Based Health Insurance<br>7= Health Insurance through Employer<br>8= Other Privately Purchased Commercial Health Insurance<br>9= Wage/income<br>10= Loan/ Ton Tin<br>11= Sale of Assets<br>12= Gift from Relative<br>13= Savings<br>14= Other | -Categorical nominal | Paym ent metho d | GADC adapted to context specific |

| | | | | | |
|---|---|---|---|---|---|
| Q28 | How much in total was spent on transport to go to and return from the selected place(s) in Q24?<br><br>*-Record 99 if don't know and 88 if refuse* | 0 = free/no cost<br>1 = in kind<br>2 = ________Riels | -Categorical nominal<br>-Quantitative Continuous | Cost of transport | CDHS |
| Q29 | On average how many hours do you spend to get treatment/advices from the selected place(s) in Q24?<br><br>*-Record 99 if don't know and 88 if refuse* | ________Hours | Quantitative continuous | Time spending | GACD book (Adapted) |
| Q30 | How satisfied are you with the effect of your {treatment/care} at the selected place(s) in Q24?<br><br>*-Record 99 if don't know/unsure* | 0 = Very satisfied<br>1 = Satisfied<br>2 = Neither satisfied nor dissatisfied<br>3 = Dissatisfied<br>4 = Very dissatisfied | Categorical ordinal | Patient satisfaction of care services | SAPS 2006 |
| Q31 | How satisfied are you with the explanations the {doctor/other health professional} has given you about the results of your {treatment/care} at the selected place(s) in Q24?<br><br>*-Record 99 if don't know/unsure* | 0 = Very satisfied<br>1 = Satisfied<br>2 = Neither satisfied nor dissatisfied<br>3 = Dissatisfied<br>4 = Very dissatisfied | Categorical ordinal | Patient satisfaction of care services | SAPS 2006 |
| Q32 | The {doctor/other health professional} at the selected place(s) in Q24 was very careful to check everything when examining you.<br><br>*-Record 99 if don't know/unsure* | 0 = Strongly agree<br>1 = Agree<br>2 = Not sure<br>3 = Disagree<br>4 = Strongly disagree | Categorical ordinal | Patient satisfaction of care services | SAPS 2006 |

| | | | | | |
|---|---|---|---|---|---|
| Q33 | At the selected place(s) in Q24, how satisfied were you with the choices you had in decisions affecting your health care?<br><br>*-Record 99 if don't know/unsure* | 0 = Very satisfied<br>1 = Satisfied<br>2 = Neither s atisfied nor dissatisfied<br>3 = Dissatisfied<br>4 = Very dissatisfied | Categorical ordinal | Patient satisfaction of care services | SAPS 2006 |
| Q34 | How much of the time did you feel respected by the {doctor/other health professional} at the selected place(s) in Q24?<br><br>*-Record 99 if don't know/unsure* | 0 = All of the time<br>1 = Most of the time<br>2 = About half the time<br>3 = Some of the time<br>4 = None of the time | Categorical ordinal | Patient satisfaction of care services | SAPS 2006 |
| Q35 | At the selected place(s) in Q24, the time you had with the {doctor/other health professional} was too short.<br><br>*-Record 99 if don't know/unsure* | 0 = Strongly agree<br>1 = Agree<br>2 = Not sure<br>3 = Disagree<br>4 = Strongly disagree | Categorical ordinal | Patient satisfaction of care services | SAPS 2006 |
| Q36 | Are you satisfied with the care you received in the selected place(s) in Q24?<br><br>*-Record 99 if don't know/unsure* | 0 = Very satisfied<br>1 = Satisfied<br>2 = Neither s atisfied nor dissatisfied<br>3 = Dissatisfied<br>4 = Very dissatisfied | Categorical ordinal | Patient satisfaction of care services | SAPS 2006 |
| Q37 | Did you get your blood pressure measured at the selected place(s) in Q24?<br><br>*-Record 99 if don't know/unsure* | 0 = No<br>1 = Yes | Categorical variable | Access to blood pressure testing | Opinion |

| | | | | | |
|---|---|---|---|---|---|
| Q38 | Did you get your blood glucose tested at the selected place(s) in Q24?<br><br>-*Record 99 if don't know/unsure* | 0 = No<br>1 = Yes | Categorical variable | Access to blood glucose testing | Opinion |
| Q39 | Would you recommend the selected place(s) in Q24 to others?<br><br>-*Record 99 if don't know/unsure* | 1 = Not recommended<br>2 = Recommend with reservations<br>3 = Recommend<br>4 = Highly recommend | Categorical ordinal | Patient satisfaction of care services | Opinion |

| SECTION 3c: HEALTH CARE UTILIZATION FOR DIABETES AND HYPERTENSION | | | | | |
|---|---|---|---|---|---|
| **Q.N** | **Description and Questions** | **Response** | **Type of Variable** | **Field** | **Ref:** |
| Q43c | How long have you lived with diabetes?<br><br>*-Record 99 if don't know/unsure and 88 if refuse*<br>*-Less than a year is rounded up to one year*<br>*-Standard rounded up formula is applied.* | ________Years | Quantitative concrete | Duration DM and HT | Opinion |
| Q44c | Where were you first diagnosed as having diabetes?<br><br>*-Record 99 if don't know and 88 if refuse* | 1= National hospital (PP)<br>2= Provincial hospital (RH)<br>3= District hospital (RH)<br>4= Health centre<br>5= Health post<br>6= Provincial rehabilitation centre (PRC) or Community-based rehabilitation (CBR)<br>7= Other public; specify:<br>8= Private hospital<br>9= Private clinic<br>10= Private pharmacy<br>11= Home/Office of trained health worker/nurse<br>12= Visit of trained health worker/nurse<br>13= Other private medical; specify:<br>14= Shop selling drugs/market<br>15= Kru Khmer/ Magician | Categorical Nominal | Diagnosis DM (CoC) | Opinion |

| | | | | | |
|---|---|---|---|---|---|
| | | 16= Monk/religious leader<br>17= Traditional birth attendant<br>18= Oversee medical service<br>19= MoPoTsyo<br>20= Other; specify: | | | |
| Q45c | Where did you first seek advice or treatment for diabetes after being diagnosed?<br><br>*-Record 99 if don't know and 88 if refuse* | 1= National hospital (PP)<br>2= Provincial hospital (RH)<br>3= District hospital (RH)<br>4= Health centre<br>5= Health post<br>6= Provincial rehabilitation centre (PRC) or Community-based rehabilitation (CBR)<br>7= Other public; specify:<br>8= Private hospital<br>9= Private clinic<br>10= Private pharmacy<br>11= Home/Office of trained health worker/nurse<br>12= Visit of trained health worker/nurse<br>13= Other private medical; specify:<br>14= Shop selling drugs/market<br>15= Kru Khmer/ Magician<br>16= Monk/religious leader<br>17= Traditional birth attendant | Categorical Nominal | Link to care DM (CoC) | CDHS (Adapted for disease specific) |

| | | | | | |
|---|---|---|---|---|---|
| | | 18= Oversee medical service<br>19= MoPoTsyo<br>20= Other; specify: | | | |
| Q46c | How long have you lived with hypertension?<br><br>*-Record 99 if don't know/unsure and 88 if refuse*<br>*-Less than a year is rounded up to one year*<br>*-Standard rounded up formula is applied.* | ________Years | Quantitative concrete | Duration DM and HT | Opinion |
| Q47c | Where were you first diagnosed as having hypertension?<br><br>*-Record 99 if don't know and 88 if refuse* | 1= National hospital (PP)<br>2= Provincial hospital (RH)<br>3= District hospital (RH)<br>4= Health centre<br>5= Health post<br>6= Provincial rehabilitation centre (PRC) or Community-based rehabilitation (CBR)<br>7= Other public; specify:<br>8= Private hospital<br>9= Private clinic<br>10= Private pharmacy<br>11= Home/Office of trained health worker/nurse<br>12= Visit of trained health worker/nurse<br>13= Other private medical; specify:<br>14= Shop selling drugs/market | Categorical Nominal | Diagnosis HT (CoC) | Opinion |

| | | | | | |
|---|---|---|---|---|---|
| | | 15= Kru Khmer/ Magician<br>16= Monk/religious leader<br>17= Traditional birth attendant<br>18= Oversee medical service<br>19= MoPoTsyo<br>20= Other; specify: | | | |
| Q48c | Where did you first seek advice or treatment for hypertension after being diagnosed?<br><br>-*Record 99 if don't know and 88 if refuse* | 1= National hospital (PP)<br>2= Provincial hospital (RH)<br>3= District hospital (RH)<br>4= Health centre<br>5= Health post<br>6= Provincial rehabilitation centre (PRC) or Community-based rehabilitation (CBR)<br>7= Other public; specify:<br>8= Private hospital<br>9= Private clinic<br>10= Private pharmacy<br>11= Home/Office of trained health worker/nurse<br>12= Visit of trained health worker/nurse<br>13= Other private medical; specify:<br>14= Shop selling drugs/market<br>15= Kru Khmer/ Magician<br>16= Monk/religious leader | Categorical Nominal | Link to care HT (CoC) | CDHS (Adapted for disease specific) |

| | | | | | |
|---|---|---|---|---|---|
| | | 17= Traditional birth attendant<br>18= Oversee medical service<br>19= MoPoTsyo<br>20= Other; specify: | | | |
| Q49c | Did you go to other places for follow up treatment/care for your hypertensive and diabetes conditions?<br><br>*-Record 88 if refuse*<br>*-If NO, please skip Q50c* | 0 = No<br>1 = Yes | Categorical binary | Trust DM and HT | CDHS |
| Q50c | If yes to Q49c, where else did you go to get follow up treatment/care for both conditions? | 1= National hospital (PP)<br>2= Provincial hospital (RH)<br>3= District hospital (RH)<br>4= Health centre<br>5= Health post<br>6= Provincial rehabilitation centre (PRC) or Community-based rehabilitation (CBR)<br>7= Other public; specify:<br>8= Private hospital<br>9= Private clinic<br>10= Private pharmacy<br>11= Home/Office of trained health worker/nurse<br>12= Visit of trained health worker/nurse<br>13= Other private medical; specify:<br>14= Shop selling drugs/market | Categorical Nominal | Link to care DM and HT (CoC) | CDHS (Adapted for disease specific) |

| | | | | | |
|---|---|---|---|---|---|
| | | 15= Kru Khmer/ Magician<br>16= Monk/religious leader<br>17= Traditional birth attendant<br>18= Oversee medical service<br>19= MoPoTsyo<br>20= Other; specify: | | | |
| Q51c | Did you get treatment/care for both conditions in the past 12 months?<br><br>*-Record 88 if refuse*<br>*-If NO, please skip Q52c-69c* | 0 = No<br>1 = Yes | Categorical binary | Retain in care DM and HT (CoC) | CDHS (Adapted for disease specific) |
| Q52c | Are you currently receiving any of the following treatment/advices for both conditions prescribed by a doctor or other health care worker?<br><br>Q. 52c.1. Insulin [ ____ ]<br>Q. 52c.2. Drugs (medication) that you have taken in the past two weeks [ ____ ]<br>Q. 52c.3. Special prescribed diet [ ____ ]<br>Q. 52c.4. Advice or treatment to lose weight [ ____ ]<br>Q. 52c.5. Advice or treatment to stop smoking<br>Q. 52c.6. Advice to reduce salt intake<br>Q. 52c.7. Advice to start or do more physical exercise | 0 = No 1= Yes | Categorical ordinal | In treatment for DM and HT (CoC) | STEPS Survey |
| Q53c | Have you had your blood glucose measured in the past 12 months?<br><br>*-Record 99 if don't know and 88 if refuse* | 0 = No 1= Yes | Categorical ordinal | In treatment for DM (CoC) | Veerle's suggestion |

| | | | | | |
|---|---|---|---|---|---|
| Q54c | Have you had your HbA1c tested in the past 12 months?<br><br>*-Record 99 if don't know and 88 if refuse* | 0 = No 1= Yes | Categorical ordinal | In treatment for DM (CoC) | Veerle's suggestion |
| Q55c | Have you had your blood cholesterol measured in the past 12 months?<br><br>*-Record 99 if don't know and 88 if refuse* | 0 = No 1= Yes | Categorical ordinal | In treatment for HT (CoC) | Veerle's suggestion |
| Q56c | Where did you seek medical advice or treatment for your conditions in **the past 3 months?**<br><br>*-**More than one answer can be selected**.*<br>*-Data collectors can use probes to help respondents determine the types of health facilities in the Response Column.*<br>*-Record 99 if don't know and 88 if refuse*<br>*-If 21=Nowhere, go to Q70c.* | 1= National hospital (PP)<br>2= Provincial hospital (RH)<br>3= District hospital (RH)<br>4= Health centre<br>5= Health post<br>6= Provincial rehabilitation centre (PRC) or Community-based rehabilitation (CBR)<br>7= Other public; specify:<br>8= Private hospital<br>9= Private clinic<br>10= Private pharmacy<br>11= Home/Office of trained health worker/nurse<br>12= Visit of trained health worker/nurse<br>13= Other private medical; specify:<br>14= Shop selling drugs/market | Categorical Nominal | Care provider_3M | CDHS and GACD book |

| | | | | | |
|---|---|---|---|---|---|
| | | 15= Kru Khmer/ Magician<br>16= Monk/religious leader<br>17= Traditional birth attendant<br>18= Oversee medical service<br>19= MoPoTsyo<br>20= Other; specify:<br>21= Nowhere | | | |
| From Q57c-Q69c, it is a set of questions that are asked following choices selected in Q56c. If 2 or 3 choices were selected in Q56c, Q57c-Q69c would appear 2 or 3 times, accordingly. | | | | | |
| Q57c | How many times did you visit the selected place**(s)** in Q56c in the past three months? | _______ times | Quantitative discrete | Number of visits | GACD |
| Q58c | How much in total was spent on the treatment at the selected place(s) in Q56c?<br><br>*-Record 99 if don't know and 88 if refuse* | 0 = free/no cost<br>1 = in kind<br>2 = ________Riels OR ________ USD | -Categorical nominal<br>- Quantitative Continuous | Cost of treatment | CDHS |
| Q59c | How did you pay for the treatment cost at the selected place(s) in Q56c?<br><br>*-Record 99 if don't know and 88 if refuse* | 1= Health Equity Fund<br>2= Voucher<br>3= Fee Exemption<br>4= NGO<br>5= National Social Security Fund<br>6= Community-Based Health Insurance<br>7= Health Insurance through Employer<br>8= Other Privately | -Categorical nominal | Payment method | GADC adapted to context specific |

| | | | | | |
|---|---|---|---|---|---|
| | | Purchased Commercial Health Insurance<br>9= Wage/income<br>10= Loan/ Ton Tin<br>11= Sale of Assets<br>12= Gift from Relative<br>13= Savings<br>14= Other | | | |
| Q60c | How much in total was spent on transport to go to and return from the selected place(s) in Q56c?<br><br>-*Record 99 if don't know and 88 if refuse* | 0 = free/no cost<br>1 = in kind<br>2 = _________Riels<br>OR<br>_________ USD | -Categorical nominal<br>-<br>Quantitative Continuous | Cost of transp ort | CDHS |
| Q61c | On average how many hours do you spend to get treatment/advices from the selected place(s) in Q56c?<br><br>-*Record 99 if don't know and 88 if refuse* | _________Hours | Quantitative continuous | Time spendi ng | GACD book (Adapted) |
| Q62c | How satisfied are you with the effect of your {treatment/care} at the selected place(s) in Q56c?<br><br>-*Record 99 if don't know/unsure* | 0 = Very satisfied<br>1 = Satisfied<br>2 = Neither satisfied nor dissatisfied<br>3 = Dissatisfied<br>4 = Very dissatisfied | Categorical ordinal | Patien t satisfa ction of care servic e | SAPS 2006 |
| Q63c | How satisfied are you with the explanations the {doctor/other health professional} has given you about the results of your {treatment/care} at the selected place(s) in Q56c?<br><br>-*Record 99 if don't know/unsure* | 0 = Very satisfied<br>1 = Satisfied<br>2 = Neither satisfied nor dissatisfied<br>3 = Dissatisfied<br>4 = Very d issatisfied | Categorical ordinal | Patien t satisfa ction of care servic e | SAPS 2006 |

| | | | | | |
|---|---|---|---|---|---|
| Q64c | The {doctor/other health professional} at the selected place(s) in Q56c was very careful to check everything when examining you.<br><br>*-Record 99 if don't know/unsure* | 0 = Strongly agree<br>1 = Agree<br>2 = Not sure<br>3 = Disagree<br>4 = Strongly disagree | Categorical ordinal | Patient satisfaction of care service | SAPS 2006 |
| Q65c | At the selected place(s) in Q56c, how satisfied were you with the choices you had in decisions affecting your health care?<br><br>*-Record 99 if don't know/unsure* | 0 = Very satisfied<br>1 = Satisfied<br>2 = Neither satisfied nor dissatisfied<br>3 = Dissatisfied<br>4 = Very dissatisfied | Categorical ordinal | Patient satisfaction of care service | SAPS 2006 |
| Q66c | How much of the time did you feel respected by the {doctor/other health professional} at the selected place(s) in Q56c?<br><br>*-Record 99 if don't know/unsure* | 0 = All of the time<br>1 = Most of the time<br>2 = About half the time<br>3 = Some of the time<br>4 = None of the time | Categorical ordinal | Patient satisfaction of care service | SAPS 2006 |
| Q67c | At the selected place(s) in Q56c, the time you had with the {doctor/other health professional} was too short.<br><br>*-Record 99 if don't know/unsure* | 0 = Strongly agree<br>1 = Agree<br>2 = Not sure<br>3 = Disagree<br>4 = Strongly disagree | Categorical ordinal | Patient satisfaction of care service | SAPS 2006 |
| Q68c | Are you satisfied with the care you received in the selected place(s) in Q56c?<br><br>*-Record 99 if don't know/unsure* | 0 = Very satisfied<br>1 = Satisfied<br>2 = Neither satisfied nor dissatisfied<br>3 = Dissatisfied<br>4 = Very dissatisfied | Categorical ordinal | Patient satisfaction of care service | SAPS 2006 |

| | | | | | |
|---|---|---|---|---|---|
| Q69c | Would you recommend the selected place(s) in Q56c to others?<br><br>*-Record 99 if don't know/unsure* | 1 = Not recommended<br>2 = Recommend with reservations<br>3 = Recommend<br>4 = Highly recommend | Categorical ordinal | Patient satisfaction of care services | Opinion |
| Q70c | Have you ever been told by a doctor that you have eyes problems?<br><br>*-Record 99 if don't know/unsure* | 0 = No<br>1 = Yes | Quantitative discrete | Complication-DM and HT | Cambodia national guideline |
| Q71c | Have you ever been told by a doctor that you have kidney problems?<br><br>*-Record 99 if don't know/unsure* | 0 = No<br>1 = Yes | Quantitative discrete | Complication-DM and HT | Cambodia national guideline |
| Q72c | Have you ever been told by a doctor that you have lost the sensation of your peripheral membrane, such as foot, hand, arm?<br><br>*-Record 99 if don't know/unsure* | 0 = No<br>1 = Yes | Quantitative discrete | Complication-DM and HT | Cambodia national guideline |
| | In **the past 3 months**, besides seeking medical advice or treatment for your hypertensive and diabetes condition, have you sought medical treatment or advice for other illnesses or conditions?<br><br>If No, go to Section 4. | 0 = No<br>1 = Yes | | | |

| | | | | | |
|---|---|---|---|---|---|
| Q24 | Where did you seek medical advice or treatment for that illness in the past 3 months?<br><br>*-More than one answer can be selected.*<br>*-Data collectors can use probes to help respondents determine the types of health facilities in the Response Column.*<br>*-Record 99 if don't know and 88 if refuse* | 1= National hospital (PP)<br>2= Provincial hospital (RH)<br>3= District hospital (RH)<br>4= Health centre<br>5= Health post<br>6= Provincial rehabilitation centre (PRC) or Community-based rehabilitation (CBR)<br>7= Other public; specify:<br>8= Private hospital<br>9= Private clinic<br>10= Private pharmacy<br>11= Home/Office of trained health worker/nurse<br>12= Visit of trained health worker/nurse<br>13= Other private medical; specify:<br>14= Shop selling drugs/market<br>15= Kru Khmer/ Magician<br>16= Monk/religious leader<br>17= Traditional birth attendant<br>18= Oversee medical service<br>19= Other; specify: | Categorical Nominal | Care provid er_3M | CDHS and GACD book |
| From Q25-Q39, it is a set of questions that are asked following choices selected in Q24. If 2 or 3 choices were selected in Q24, Q25-Q39 would appear 2 or 3 times, accordingly. | | | | | |

| | | | | | |
|---|---|---|---|---|---|
| Q25 | How many times did you visit the selected place**(s)** in Q24 in the past three months? | _______ times | Quantitative discrete | Numb er of visits | GACD |
| Q26 | How much in total was spent on the treatment at the selected place(s) in Q24?<br><br>-*Record 99 if don't know and 88 if refuse* | 0 = free/no cost<br>1 = in kind<br>2 = ________Riels | -Categorical nominal<br>-<br>Quantitative Continuous | Cost of treatm ent | CDHS |
| Q27 | How did you pay for the treatment cost at the selected place(s) in Q24?<br><br>-*Record 99 if don't know and 88 if refuse* | 1= Health Equity Fund<br>2= Voucher<br>3= Fee Exemption<br>4= NGO<br>5= National Social Security Fund<br>6= Community-Based Health Insurance<br>7= Health Insurance through Employer<br>8= Other Privately Purchased Commercial Health Insurance<br>9= Wage/income<br>10= Loan/ Ton Tin<br>11= Sale of Assets<br>12= Gift from Relative<br>13= Savings<br>14= Other | -Categorical nominal | Paym ent metho d | GADC adapted to context specific |

| | | | | | |
|---|---|---|---|---|---|
| Q28 | How much in total was spent on transport to go to and return from the selected place(s) in Q24?<br><br>*-Record 99 if don't know and 88 if refuse* | 0 = free/no cost<br>1 = in kind<br>2 = ________Riels | -Categorical nominal<br>-Quantitative Continuous | Cost of transport | CDHS |
| Q29 | On average how many hours do you spend to get treatment/advices from the selected place(s) in Q24?<br><br>*-Record 99 if don't know and 88 if refuse* | ________Hours | Quantitative continuous | Time spending | GACD book (Adapted) |
| Q30 | How satisfied are you with the effect of your {treatment/care} at the selected place(s) in Q24?<br><br>*-Record 99 if don't know/unsure* | 0 = Very satisfied<br>1 = Satisfied<br>2 = Neither satisfied nor dissatisfied<br>3 = Dissatisfied<br>4 = Very dissatisfied | Categorical ordinal | Patient satisfaction of care services | SAPS 2006 |
| Q31 | How satisfied are you with the explanations the {doctor/other health professional} has given you about the results of your {treatment/care} at the selected place(s) in Q24?<br><br>*-Record 99 if don't know/unsure* | 0 = Very satisfied<br>1 = Satisfied<br>2 = Neither satisfied nor dissatisfied<br>3 = Dissatisfied<br>4 = Very dissatisfied | Categorical ordinal | Patient satisfaction of care services | SAPS 2006 |
| Q32 | The {doctor/other health professional} at the selected place(s) in Q24 was very careful to check everything when examining you.<br><br>*-Record 99 if don't know/unsure* | 0 = Strongly agree<br>1 = Agree<br>2 = Not sure<br>3 = Disagree<br>4 = Strongly disagree | Categorical ordinal | Patient satisfaction of care services | SAPS 2006 |

| | | | | | |
|---|---|---|---|---|---|
| Q33 | At the selected place(s) in Q24, how satisfied were you with the choices you had in decisions affecting your health care?<br><br>*-Record 99 if don't know/unsure* | 0 = Very satisfied<br>1 = Satisfied<br>2 = Neither s atisfied nor dissatisfied<br>3 = Dissatisfied<br>4 = Very dissatisfied | Categorical ordinal | Patient satisfaction of care services | SAPS 2006 |
| Q34 | How much of the time did you feel respected by the {doctor/other health professional} at the selected place(s) in Q24?<br><br>*-Record 99 if don't know/unsure* | 0 = All of the time<br>1 = Most of the time<br>2 = About half the time<br>3 = Some of the time<br>4 = None of the time | Categorical ordinal | Patient satisfaction of care services | SAPS 2006 |
| Q35 | At the selected place(s) in Q24, the time you had with the {doctor/other health professional} was too short.<br><br>*-Record 99 if don't know/unsure* | 0 = Strongly agree<br>1 = Agree<br>2 = Not sure<br>3 = Disagree<br>4 = Strongly disagree | Categorical ordinal | Patient satisfaction of care services | SAPS 2006 |
| Q36 | Are you satisfied with the care you received in the selected place(s) in Q24?<br><br>*-Record 99 if don't know/unsure* | 0 = Very satisfied<br>1 = Satisfied<br>2 = Neither s atisfied nor dissatisfied<br>3 = Dissatisfied<br>4 = Very dissatisfied | Categorical ordinal | Patient satisfaction of care services | SAPS 2006 |
| Q37 | Did you get your blood pressure measured at the selected place(s) in Q24?<br><br>*-Record 99 if don't know/unsure* | 0 = No<br>1 = Yes | Categorical variable | Access to blood pressure testing | Opinion |

| | | | | | |
|---|---|---|---|---|---|
| Q38 | Did you get your blood glucose tested at the selected place(s) in Q24?<br><br>-*Record 99 if don't know/unsure* | 0 = No<br>1 = Yes | Categorical variable | Access to blood glucose testing | Opinion |
| Q39 | Would you recommend the selected place(s) in Q24 to others?<br><br>-*Record 99 if don't know/unsure* | 1 = Not recommended<br>2 = Recommend with reservations<br>3 = Recommend<br>4 = Highly recommend | Categorical ordinal | Patient satisfaction of care services | Opinion |
