## Supplementary material for "Healthcare utilization and expenditure among people with type 2 diabetes and/or hypertension in Cambodia: results from a cross-sectional survey": Annexure 4

### Annexure 4. Arithmetic and the geometric mean of medical cost by participant characteristics in Cambodia, 2020

*Table S1. Arithmetic mean of medical cost by participant characteristics in Cambodia, 2020*

| Variable | **Overall** | **No T2D/HTN** | **HTN** | **T2D** | **T2D plus HTN** |
|---|---|---|---|---|---|
| | Mean [95% CI] | Mean [95% CI] | Mean [95% CI] | Mean [95% CI] | Mean [95% CI] |
| Sex | | | | | |
| Male | 16.8 [15.2–18.5] | 16.8 [14.7–18.9] | 15.5 [12.3–18.6] | 25.5 [15.8–35.2] | 17.0 [10.2–23.7] |
| Female | 19.5 [18.3–20.6] | 22.6 [20.8–24.4] | 14.3 [12.7–15.8] | 21.1 [16.0–26.2] | 22.2 [18.9–25.5] |
| *P*-value | 0.015 | <0.001 | 0.480 | 0.393 | 0.212 |
| Age in years | | | | | |
| 40–49 | 19.0 [16.8–21.2] | 20.6 [18.1–23.2] | 20.6 [18.1–23.2] | 10.3 [6.5–14.0] | 19.7 [7.1–32.4] |
| 50–59 | 19.1 [17.5–20.7] | 82.8 [73.0–92.6] | 20.7 [18.3–23.2] | 14.2 [12.0–16.5] | 24.5 [17.6–31.3] |
| 60+ | 18.2 [16.8–19.7] | 82.1 [72.8–91.4] | 20.5 [18.2–22.9] | 15.4 [13.4–17.4] | 21.1 [13.8–28.4] |
| *P*-value | 0.698 | 0.995 | 0.120 | 0.704 | 0.850 |
| Educational level | | | | | |
| No schooling | 19.1 [17.4–20.9] | 20.6 [17.9–23.3] | 16.2 [13.8–18.7] | 22.8 [14.8–30.7] | 22.1 [15.8–28.5] |
| Primary | 17.9 [16.7–19.1] | 20.3 [18.5–22.1] | 12.9 [11.1–14.7] | 20.5 [15.1–26.0] | 19.9 [16.7–23.2] |
| Secondary/higher | 21.1 [18.0–24.3] | 22.0 [17.9–26.2] | 16.9 [11.5–22.2] | 28.6 [8.8–48.4] | 26.8 [14.3–39.2] |
| *P*-value | 0.091 | 0.726 | 0.054 | 0.506 | 0.365 |
| Having NSSF membership | | | | | |
| No | 18.8 [17.8–19.7] | 20.7 [19.2–22.1] | 14.5 [13.1–16.0] | 22.9 [18.1–27.6] | 21.9 [18.8–25.0] |
| Yes | 17.5 [13.2–21.8] | 20.4 [13.9–26.9] | 14.7 [7.0–22.4] | 13.8 [1.6–26.0] | 11.8 [4.7–18.8] |
| *P*-value | 0.5604 | 0.937 | 0.961 | 0.339 | 0.076 |
| Having HEF membership | | | | | |
| No | 19.4 [18.3–20.5] | 21.4 [19.8–23.0] | 14.9 [13.3–16.5] | 24.1 [18.8–29.3] | 22.6 [19.4–25.9] |
| Yes | 15.8 [13.8–17.8] | 17.6 [14.6–20.5] | 13.2 [10.2–16.2] | 14.3 [11.0–18.6] | 14.9 [7.5–22.3] |
| *P*-value | 0.004 | 0.035 | 0.360 | 0.096 | 0.058 |
| Household socio-economic class | | | | | |
| Poorest | 20.2 [17.7–22.8] | 23.9 [20.1–27.6] | 13.6 [10.3–16.9] | 18.0 [6.6–29.4] | 23.6 [14.8–32.4] |
| Poor | 16.9 [14.8–18.9] | 17.8 [15.0–20.6] | 14.7 [11.4–18.0] | 23.2 [11.3–35.1] | 14.8 [6.0–23.7] |
| Medium | 18.2 [16.2–20.2] | 19.8 [16.9–22.6] | 14.7 [11.4–18.0] | 18.8 [12.5–25.0] | 21.2 [14.4–28.1] |
| Rich | 20.0 [17.8–22.2] | 23.0 [19.4–26.5] | 15.6 [12.5–18.6] | 23.4 [10.3–36.6] | 21.5 [16.8–26.3] |
| Richest | 18.2 [16.2–20.2] | 19.2 [16.4–22.0] | 14.0 [11.0–17.0] | 26.8 [15.8–37.7] | 24.1 [17.4–30.8] |
| *P*-value | 0.171 | 0.034 | 0.919 | 0.687 | 0.427 |
| Sector | | | | | |
| Private | 20.5 [19.3–21.6] | 23.0 [21.4–24.7] | 15.7 [14.0–17.3] | 24.9 [18.3–31.6] | 22.0 [18.0–26.0] |
| Public | 12.0 [10.3–13.7] | 9.8 [7.4–12.2] | 10.6 [7.7–13.6] | 18.5 [13.2–23.8] | 20.4 [15.6–25.2] |
| Both | 21.7 [17.4–26.1] | 24.8 [19.5–30.1] | 9.4 [5.7–13.1] | — | 20.5 [10.6–30.4] |

| | | | | | |
|---|---|---|---|---|---|
| *P*-value | <0.001 | <0.001 | 0.013 | 0.164 | 0.878 |
| Care model | | | | | |
| Coexisting | 16.7 [14.7–18.7] | 17.9 [15.1–20.6] | 13.8 [10.8–16.9] | 20.0 [11.8–28.2] | 23.3 [12.1–34.6] |
| Community-based | 22.6 [20.3–25.0] | 24.5 [21.1–27.9] | 18.9 [15.1–22.7] | 22.3 [12.7–32.0] | 27.2 [19.2–35.2] |
| Health center-based (high) | 18.6 [16.4–20.7] | 20.7 [17.5–24.0] | 14.8 [11.7–17.8] | 19.7 [9.2–30.1] | 22.8 [14.6–30.9] |
| Health center-based (low) | 15.7 [13.9–17.5] | 17.6 [14.8–20.4] | 11.0 [8.8–13.2] | 24.1 [13.1–35.2] | 19.0 [15.1–22.9] |
| Hospital-based | 20.3 [17.9–22.7] | 22.8 [19.4–26.2] | 14.8 [10.9–18.7] | 23.3 [12.1–34.6] | 18.1 [12.0–24.1] |
| *P*-value | <0.001 | 0.006 | 0.011 | 0.896 | 0.2695 |

*Abbreviation CI, confidence interval*

*Note: Health center-based (high) means the OD with high coverage (six out of nine) of health centers with the WHO PEN; Health center-based (low) means the OD with low coverage (six out of 25) of health centers with the WHO PEN*

*Table S2. Geometric mean of medical cost by participant characteristics in Cambodia, 2020*

| Variable | **Overall** | **No T2D/HTN** | **HTN** | **T2D** | **T2D plus HTN** |
|---|---|---|---|---|---|
| | Mean [95% CI] | Mean [95% CI] | Mean [95% CI] | Mean [95% CI] | Mean [95% CI] |
| Sex | | | | | |
| Male | 6.9 [6.2–7.8] | 6.5 [5.6–7.5] | 7.0 [5.7–8.6] | 14.7 [8.8–24.7] | 8.5 [4.2–17.0] |
| Female | 8.5 [7.9–9.1] | 8.9 [8.0–10.0] | 6.8 [6.1–7.6] | 12.5 [9.4–16.5] | 13.9 [11.1–17.5] |
| *P*-value | **0.004** | **<0.001** | 0.79 | 0.548 | **0.104** |
| Age in years | | | | | |
| 40–49 | 7.9 [6.8–9.0] | 8.2 [7.0–9.7] | 8.2 [7.0–9.7] | 5.0 [3.7–6.8] | 13.2 [8.3–20.9] |
| 50–59 | 8.4 [7.6–9.4] | 28.2 [23.6–33.6] | 7.9 [6.7–9.2] | 7.6 [6.4–8.9] | 15.2 [10.6–21.9] |
| 60+ | 7.7 [7.0–8.5] | 28.4 [23.7–33.9] | 7.9 [6.8–9.3] | 6.9 [6.0–7.8] | 11.1 [7.1–17.3] |
| *P*-value | 0.480 | 0.91 | **0.09** | 0.509 | 0.384 |
| Educational level | | | | | |
| No schooling | 7.9 [7.0–8.8] | 7.4 [6.2–8.8] | 7.6 [6.4–8.9] | 13.0 [8.1–20.8] | 11.6 [7.4–18.2] |
| Primary | 8.0 [7.4–8.7] | 8.3 [7.4–9.4] | 6.4 [5.7–7.3] | 12.6 [9.2–17.3] | 12.9 [9.8–17.0] |
| Secondary/higher | 8.2 [6.8–9.9] | 8.0 [6.2–10.4] | 6.6 [4.7–9.4] | 15.7 [7.0–35.1] | 17.2 [9.2–32.2] |
| *P*-value | 0.920 | 0.510 | 0.299 | 0.842 | 0.600 |
| Having NSSF membership | | | | | |
| No | 8.1 [7.6–8.6] | 8.0 [7.3–8.8] | 7.0 [6.3–7.7] | 13.6 [10.6–17.4] | 13.5 [10.8–16.9] |
| Yes | 6.8 [5.0–9.2] | 7.9 [5.2–12.0] | 5.3 [3.0–9.2] | 7.6 [1.5–37.8] | 6.3 [2.2–18.1] |
| *P*-value | **0.2351** | 0.937 | 0.234 | 0.261 | **0.071** |
| Having HEF membership | | | | | |
| No | 8.8 [8.2–9.4] | 8.7 [7.9–9.7] | 7.4 [6.6–8.2] | 14.1 [7.4–20.9] | 16.1 [13.5–19.3] |
| Yes | 5.4 [4.6–6.3] | 5.5 [4.4–6.9] | 5.1 [4.0–6.7] | 8.6 [4.5–16.3] | 4.1 [1.7–9.9] |
| *P*-value | **<0.001** | **<0.001** | **0.004** | **0.114** | **<0.001** |
| Household socio-economic class | | | | | |

| | | | | | |
|---|---|---|---|---|---|
| Poorest | 6.8 [5.8–8.0] | 7.3 [5.8–9.2] | 5.4 [4.1–6.9] | 8.4 [3.9–18.3] | 11.1 [5.5–22.4] |
| Poor | 7.4 [6.5–8.5] | 7.4 [6.1–8.9] | 7.1 [5.7–8.8] | 13.6 [7.2–25.9] | 6.6 [3.2–13.7] |
| Medium | 8.4 [7.3–9.6] | 8.2 [6.7–10.0] | 7.4 [6.0–9.1] | 12.6 [7.9–20.1] | 12.5 [7.5–20.8] |
| Rich | 9.4 [8.2–10.8] | 9.6 [7.8–11.9] | 7.9 [6.5–9.6] | 14.1 [6.8–28.9] | 15.8 [11.1–22.5] |
| Richest | 8.2 [7.2–9.3] | 7.8 [6.5–9.4] | 6.7 [5.5–8.3] | 16.5 [10.5–25.7] | 19.0 [14.4–25.1] |
| *P*-value | **0.022** | 0.366 | **0.133** | 0.538 | **0.054** |
| Sector | | | | | |
| Private | 9.8 [9.2–10.5] | 10.2 [9.2–11.2] | 8.4 [7.6–9.2] | 15.0 [11.2–20.2] | 15.5 [12.6–19.0] |
| Public | 3.5 [3.0–4.1] | 2.5 [2.0–3.1] | 3.1 [2.4–4.1] | 10.7 [7.0–16.3] | 9.5 [5.9–15.2] |
| Both | 13.9 [10.8–17.9] | 16.2 [12.2–21.5] | 6.9 [4.0–12.1] | — | 20.3 [12.7–32.4] |
| *P*-value | **<0.001** | **<0.001** | **<0.001** | **0.176** | **0.084** |
| Care model | | | | | |
| Coexisting | 7.3 [6.3–8.5] | 7.5 [6.2–9.2] | 6.0 [4.7–7.6] | 12.8 [6.9–23.5] | 17.5 [10.8–28.3] |
| Community-based | 10.8 [9.4–12.3] | 10.4 [8.5–12.8] | 9.7 [7.9–11.8] | 16.5 [10.8–25.2] | 20.5 [14.3–29.2] |
| Health center-based (high) | 8.5 [7.5–9.7] | 8.6 [7.0–10.5] | 7.4 [6.1–9.0] | 11.4 [7.1–18.2] | 15.0 [9.5–23.9] |
| Health center-based (low) | 6.4 [5.6–7.3] | 5.9 [4.8–7.2] | 5.6 [4.6–6.9] | 13.7 [6.9–27.4] | 13.9 [9.5–20.1] |
| Hospital-based | 7.9 [6.7–9.2] | 8.5 [6.9–10.5] | 6.3 [4.8–8.1] | 17.5 [10.8–28.3] | 7.3 [4.2–12.8] |
| *P*-value | **<0.001** | **0.002** | **0.003** | 0.911 | **0.015** |

*Abbreviation CI, confidence interval*

*Note: Health center-based (high) means the OD with high coverage (six out of nine) of health centers with the WHO PEN; Health center-based (low) means the OD with low coverage (six out of 25) of health centers with the WHO PEN*
